# Diffusion Radiomic Features of the Language Network Predict the Conversion from Mild Cognitive Impairment to Alzheimer’s

**DOI:** 10.64898/2026.01.29.26345111

**Authors:** Fatemeh Jamshidian, Mohadeseh Hosseini, Mina Kiani, Farhad Zarei, Ramtin Sanjabi, Samira Raminfard

## Abstract

**Background:** Mild cognitive impairment (MCI) precedes Alzheimer’s disease (AD) in ∼40% of cases, with early language deficits distinguishing converters. This study develops a DTI radiomics model from language network gray matter to predict MCI to AD conversion and identify preclinical biomarkers.

**Methods:** This retrospective case-control study analyzed diffusion tensor imaging (DTI) data from 97 individuals with MCI (29 converters, 68 non-converters) from the Alzheimer’s Disease Neuroimaging Initiative (ADNI). Ethical approval and participant consent were obtained by ADNI. Radiomic features were extracted from fractional anisotropy (FA) and mean diffusivity (MD) maps within language network gray matter. A logistic regression model using eleven selected features performed classification. Performance was evaluated using area under the receiver operating characteristic curve (AUC). Radiomic–cognitive associations were analyzed using Pearson correlations; group differences were assessed with Fisher’s r-to-z transformation.

**Results:** The model achieved cross-validation AUC = 0.84 and test AUC = 0.83. SHAP analysis identified two top predictors: lower right temporal pole original_glcm_Correlation_FA and higher right frontal orbital cortex original_glszm_SmallAreaHighGrayLevelEmphasis_FA. Right frontal orbital cortex original_glszm_SmallAreaHighGrayLevelEmphasis_FA correlated positively with ADAS-Q4 in non-converters (r = 0.27, p < 0.001) but negatively in converters (r = –0.48, p < 0.001).

**Conclusions:** A DTI radiomics model achieved AUC = 0.83 for predicting MCI to AD conversion, with bilateral language network microstructural features showing group-specific cognitive associations, supporting their potential as early Alzheimer’s risk biomarkers.

**Key points:**

- New method identifies Alzheimer’s risk before significant cognitive decline occurs
- Brain language regions show detectable changes in future converters
- Texture analysis reveals early disease signatures in brain tissue

## Background

Alzheimer’s disease (AD) is the most common progressive neurodegenerative disease, consisting of more advanced cognitive deficits of memory and executive and linguistic function loss, and significantly interrupts daily life. Mild Cognitive Impairment (MCI) is the initial stage of AD, a type of dementia. MCI is a cognitive dysfunction without loss of daily function (Wang et al., 2024). MCI is found to be roughly 40% more prone to convert to AD compared with normal people (Skolariki et al., 2021). Therefore, the detection of brain changes at this stage is extremely necessary in order to create new therapies.

MCI patients present with specific deficits in language compared to normal adults, but the deficits are less severe compared to AD. Language disorders could therefore differentiate MCI individuals at risk of converting into AD from those who are not (Skolariki et al., 2021). Advanced MRI methods like diffusion tensor imaging (DTI) have been beneficial in distinguishing MCI patients from AD ones. This is especially true when using two diffusion parameters: fractional anisotropy (FA) and mean diffusivity (MD) (Chandra et al., 2019). These microstructural changes could, in effect, be used to differentiate between converted and non-converted MCI (Bruggen et al., 2012). Radiomics is a new method of improving the prospective prediction of the conversion of MCI into AD disease.

Radiomics is a method that extracts many quantitative features from medical images (Kumar et al., 2012). The method enhances the effectiveness of distinguishing the converted MCI from the non-converted MCI and discovers information hidden from the regular analysis (Zhou et al., 2019). In the present study, we assessed the radiomic features extracted from the FA and MD of the language network. We developed a model of AD progression prediction with the aid of radiomics and also a model that identifies corresponding imaging biomarkers.

## Methods

### Participants

The Alzheimer’s Disease Neuroimaging Initiative (ADNI) database (adni.loni.usc.edu), accessed in May 2024, provided all of the data used in this study. ADNI is a large-scale, multimodal research initiative launched in 2003 as a $60 million, 5-year public-private partnership funded by the National Institute on Aging, the National Institute of Biomedical Imaging and Bioengineering, the Food and Drug Administration, private pharmaceutical companies, and non-profit organizations. After extracting data, quality control was conducted on the obtained information. Cases with missing clinical information or imaging data were excluded, and 97 MCI patients were included. We selected MCI subjects who converted to Alzheimer’s disease within 1 to 5 years of follow-up (n = 29; converted group), and those who remained MCI during the same period (n = 68; non-converted group). These patients had complete clinical and imaging information, including sex, age, race, education level, marital status, APOE4 gene status, cognitive test score, and DTI images. Following data extraction, quality control procedures were applied.

### Data acquisition

This study utilized data from the ADNI database (http://www.loni.ucla.edu/ADNI/). We specifically selected participants diagnosed with MCI who had DTI during ADNI phases 1, GO/2, and 3. The DTI scans were acquired using either a 3.0 Tesla GE Medical System or a Siemens scanner, both employing identical acquisition parameters. These parameters included a 256 × 256 pixel matrix, a pixel size of 1.4 mm, and a slice thickness of 2.7 mm. An Echo Planar/Spin Echo (EP/SE) pulse sequence was used, with a repetition time (TR) of 13,000 ms, an echo time (TE) of 68.3 ms, and a flip angle of 90 degrees. Diffusion encoding was performed with 31 directions, 30 diffusion-weighted images with a b-value of 1000 s/mm², and one non-diffusion-weighted (b0) image.

### Pre-processing

The DTI data was processed and analyzed using the ExploreDTI software (https://www.ExploreDTI.com) within MATLAB R2022b. This preprocessing involved correcting for eddy current-induced echo planar imaging (EPI) distortions to minimize signal artifacts. DTI scans were normalized to the MNI 152 template. FA and MD were then extracted to assess diffusion anisotropy and overall diffusion.

### Segmentation and Feature Extraction

In order to obtain segmentations of the language network, we chose ten anatomically and functionally relevant brain areas from the Harvard-Oxford Probabilistic Atlas(Desikan et al., 2006). These brain areas are well-known to consist of major components of the language network and span the frontal, temporal, and parietal lobes. Specifically, they include the Angular Gyrus, Frontal Orbital Cortex, Inferior Frontal Gyrus (pars opercularis and pars triangularis), Middle Temporal Gyrus (posterior division), Planum Temporale, Superior Frontal Gyrus, Superior Temporal Gyrus (anterior and posterior divisions), Supramarginal Gyrus (posterior division), and the Temporal Pole. These regions support major aspects of linguistic processing that involve semantic integration, phonological processing, syntactic comprehension, and speech production. The network ROIs were segregated into left and right hemispheres to control for typical asymmetry in the brain region supporting language. These masks were then applied to FA and MD maps from diffusion MRI. We then quantified and segmented 93 features per ROI map using Pyradiomics (V3.0.1) (van Griethuysen et al., 2017). In total, 4092 features were processed to capture complex tissue patterns and enhance AD prediction. This improved model performance.

These features, which are typically classified into eight categories, include a first-order statistical feature which represents the distribution of voxel intensity in the picture area defined by the mask using common and basic measures. These measures include variance, mean, and median. A shape (3D) feature that is unaffected by the gray-level intensity distribution and has information on the ROI’s three-dimensional size and shape.

This feature’s measurements also include volume, surface area, and compactness. The gray level intensity distribution has no effect on shape-based (2D) features, which quantify the ROI’s two-dimensional geometric aspects such as area, perimeter, and density.

Other features include the Gray Level Coherence Matrix (GLCM) features, which measure the spatial relationship between pixel intensities to capture texture patterns such as contrast, correlation, and homogeneity, and provide a second-order statistical analysis; the Gray Level Area Size Matrix (GLSZM) features, which describe the distribution of contiguous areas with the same intensity in the ROI, reflecting texture uniformity and heterogeneity; the Gray Level Run Length Matrix (GLRLM) features, which measure the distance between consecutive pixels with the same intensity to provide a sense of texture granularity and image patterns; the Gray Level Neighbor Color Difference Matrix (NGTDM) features, which evaluate the intensity difference between neighboring pixels, revealing aspects of image roughness and texture complexity; and finally the Gray Level Dependency Matrix (GLDM) features, which capture the spatial dependence of pixel intensities and showing how intensities within an image are related to each other at different distances, as well as the complexity.

### Feature Selection and Classification

We first preprocessed the data to ensure quality and consistency. This involved removing duplicate columns, assessing feature skewness, and handling outliers. We normalized the training data to reduce variability in radiomics feature scales using the StandardScaler method. This ensured balanced input for feature selection. Test group features were normalized using the same method as the training group.

We used a two-step approach to reduce feature dimensionality. The Maximum Relevance Minimum Redundancy (mRMR) algorithm was first applied to remove redundant features. Second, we applied a 5-fold cross-validation with the Least Absolute Shrinkage and Selection Operator (LASSO) for the final feature selection.

We establish a logistic regression classification model with a 5-fold cross-validation for the classification task. Receiver operating characteristic (ROC) curves and the corresponding areas under the curve (AUC) were utilized to assess the diagnostic capabilities of the radiomics features. The flow chart of the radiomic analysis was shown in Figure 1.

**Figure 1.**
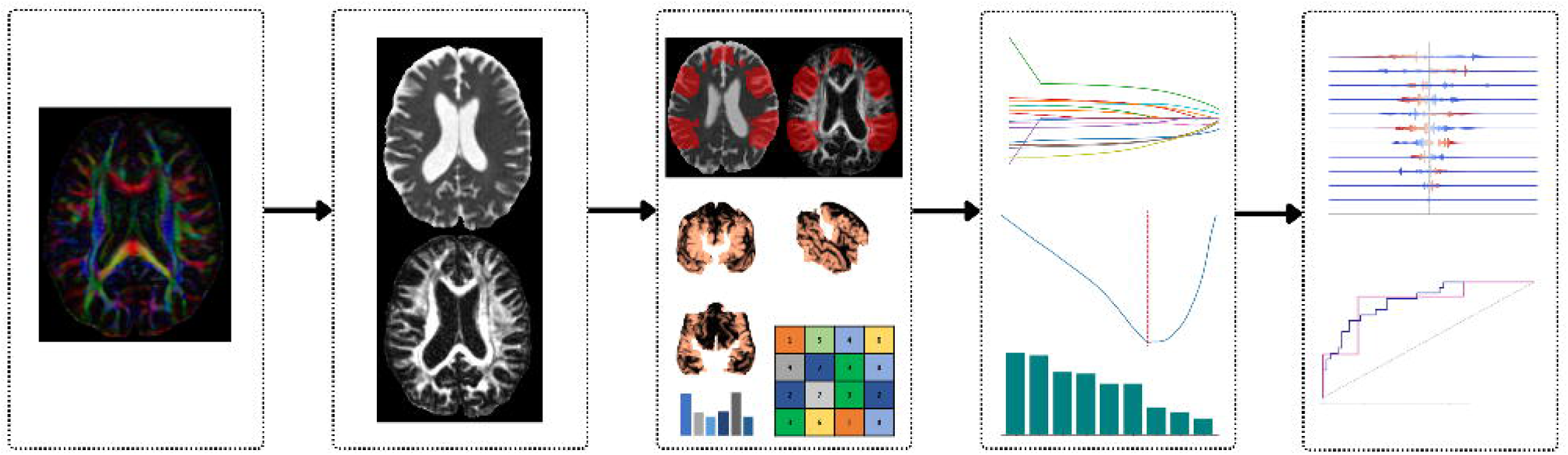
Radiomic Model Workflow

### Statistical Analysis

Statistical analyses were conducted using Python (v3.10, SciPy library) and IBM SPSS Statistics (version 27). Continuous variables were reported as mean ± standard deviation. Normality was assessed using the Shapiro–Wilk test. Independent two-sample t-tests were used for normally distributed variables, while Mann–Whitney U tests were applied for non-normally distributed variables. Categorical variables were compared using the chi-square test. A p-value < 0.05 was considered statistically significant.

## Results

### Demographic and Clinical Characteristics

The patient’s detailed demographic and clinical characteristics are presented in Table 1. The Montreal Cognitive Assessment (MoCA) is a brief test of language and other cognitive skills. It measures naming, sentence repetition, and verbal fluency. Lower scores reflect poorer language ability. The Alzheimer’s Disease Assessment Scale – Cognitive Subscale, Question 4 (ADAS-Q4) measures word recognition, assessing language indirectly through word memory, and higher scores show greater deficits. The Alzheimer’s Disease Assessment Scale – Cognitive Subscale, 11-item version (ADAS-11) evaluates language through naming, command-following, and word-finding tasks. Higher scores indicate reduced performance. The Alzheimer’s Disease Assessment Scale – Cognitive Subscale, 13-item version (ADAS-13) adds memory measures to the ADAS-11 language tasks, and higher scores correspond to more severe language and cognitive impairment. The ADASQ4, ADAS11, and ADAS13 scores of the converted group were higher than those of the non-converted group, and the MOCA score was significantly lower than that of the non-converted group (*p* < 0.001), which is consistent with previous suggests that language function is more severely impaired in MCI patients who progress to AD compared to those who remain stable. There were no significant differences in gender, age, EcogSPLang, or EcogPtLang among the two groups (in all, *p* > 0.05).

**Table 1.** The Clinical and Demographic Characteristics Of Subjects.

| *Characteristic* | *Non-Converted (n = 68)* | *Converted (n = 29)* | *p-value* |
| --- | --- | --- | --- |
| *Sex (M/F)* | 41 / 27 | 17 / 12 | 0.87 |
| *Age (years)* | 73.11 ± 7.35 | 74.96 ± 6.38 | 0.803 |
| *MOCA* | 24.37 ± 2.68 | 21.86 ± 3.03 | <0.001 |
| *ADASQ4* | 4.41 ± 2.031 | 6.72 ± 2.051 | <0.001 |
| *ADAS11* | 8.36 ± 3.93 | 11.85 ± 5.12 | 0.002 |
| *ADAS13* | 13.27 ± 5.47 | 19.41 ± 6.66 | <0.001 |
| *EcogPtLang* | 1.92 ± 0.68 | 1.93 ± 0.76 | 0.65 |
| *EcogSPLang* | 1.50 ± 0.48 | 1.94 ± 0.82 | 0.04 |

### Radiomic Feature Extraction and Selection

Radiomic features were extracted from diffusion-derived maps (FA and MD) for each subject. An initial set of 4,092 features was computed per subject. After preprocessing, including normalization and removal of low-variance or highly correlated features, 4,045 features were retained for further analysis.

To reduce dimensionality and retain the most informative features, a two-step feature selection process was applied. First, mRMR was used to select the top 15 features. Next, LASSO was employed with the optimal regularization parameter (λ), identified through five-fold cross-validation (Figure 2). This resulted in a final subset of 11 features (Table 2).

**Figure 2.**
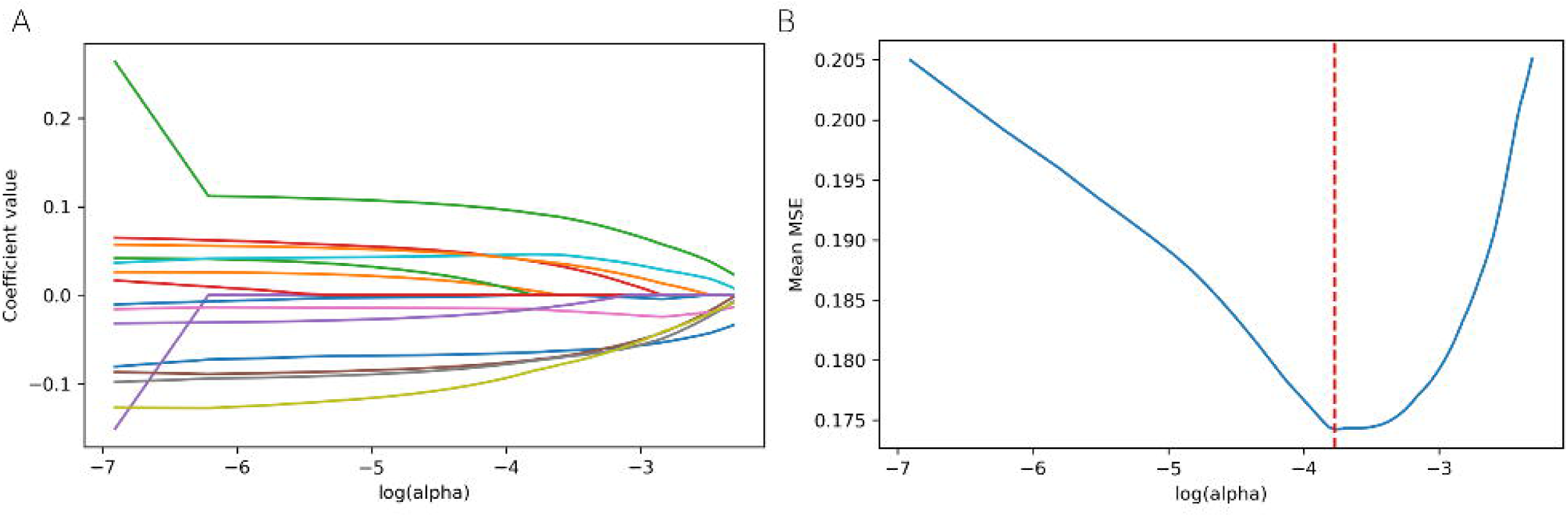
Radiomic feature selection using the LASSO method. · (A) The model fitting performance under various log(λ) values is shown in this picture. (B), This figure shows how feature coefficients vary with log(λ) values.

**Table 2.**
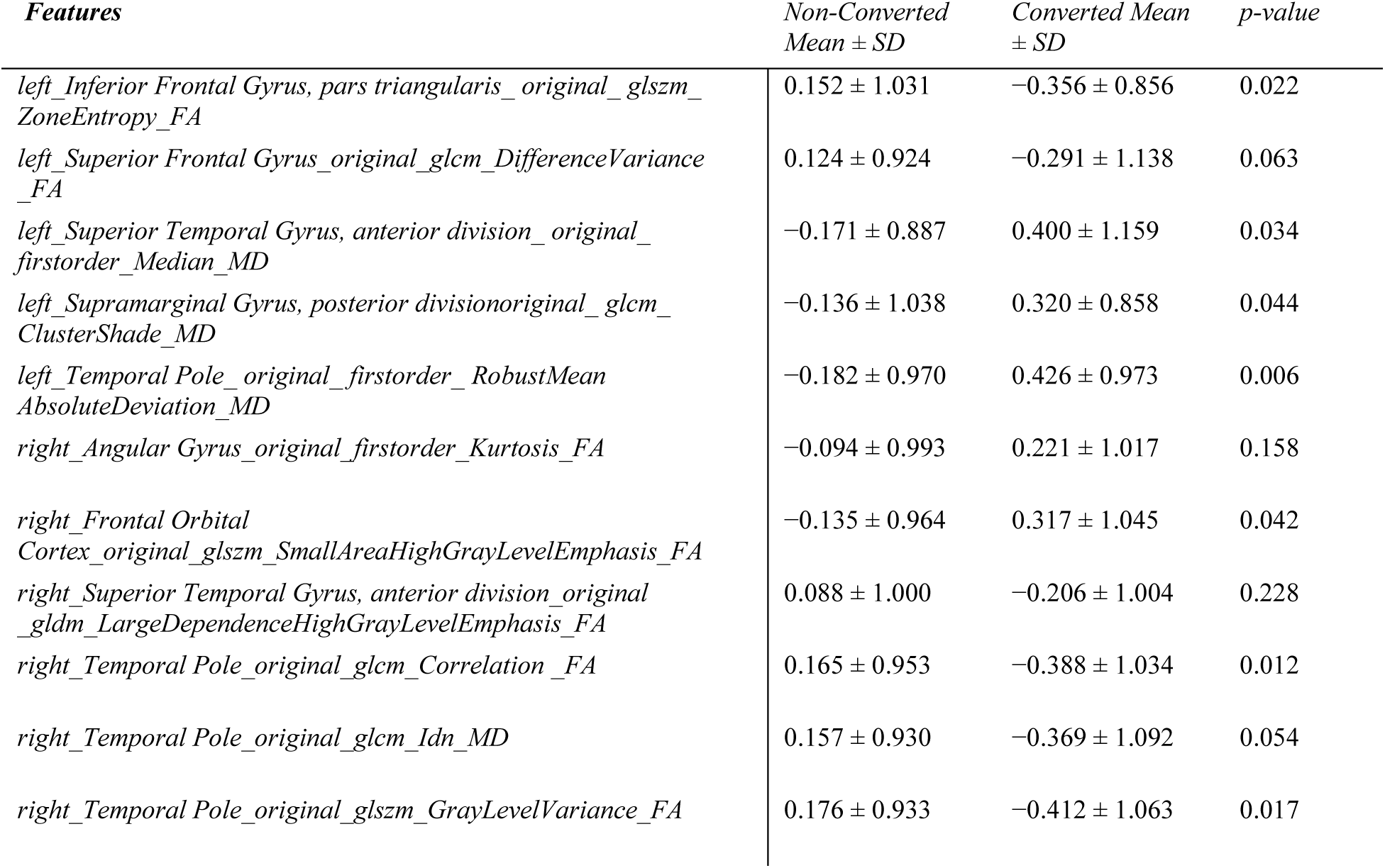
Radiomic features derived from FA and MD maps selected through a two-step feature selection procedure (mRMR + LASSO) for classification.

### Model development and Evaluation

To develop the predictive model, the training data were stratified by diagnosis and split into five folds. Each fold was iteratively used as a test set while the remaining folds were used for training and internal validation. A logistic regression classifier was trained in each fold using five-fold cross-validation.

The radiomic model demonstrated strong classification performance. The AUC during cross-validation was 0.84. When evaluated on the held-out test set (unseen data), the model achieved an AUC of 0.83, indicating robust generalization performance (Figure 3).

**Figure 3.**
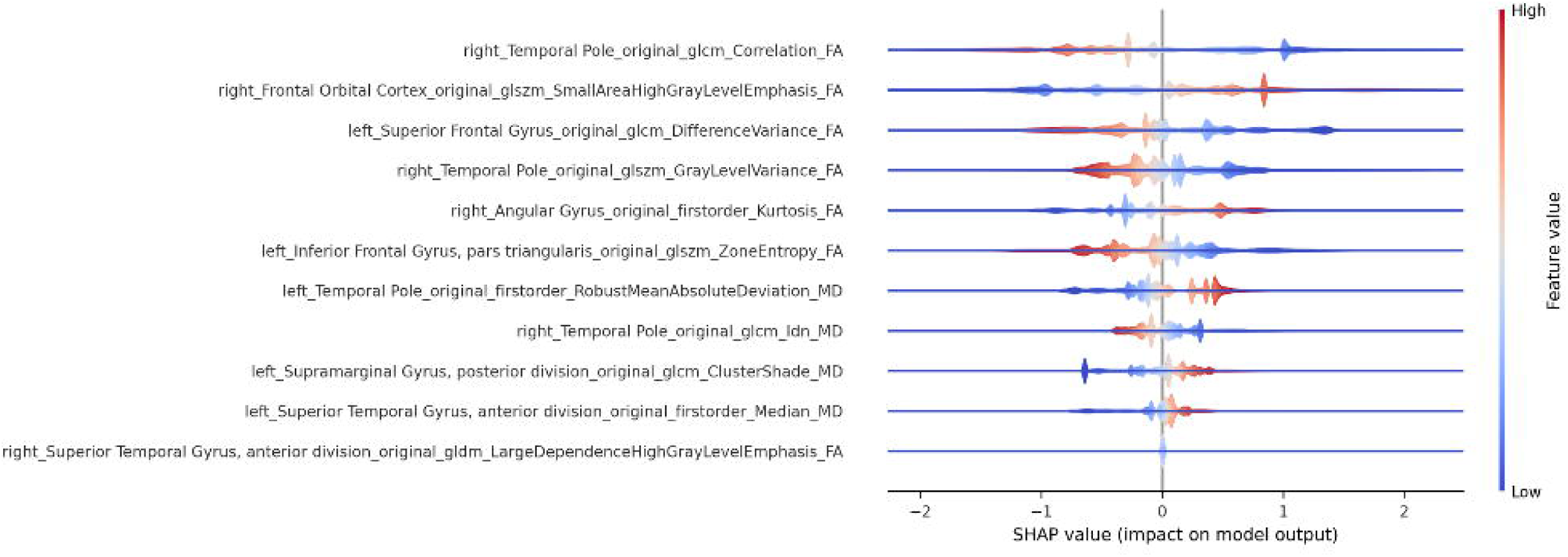
Evaluating the model’s performance on the test and train sets

To interpret the model and quantify the contribution of individual features, we applied SHAP (SHapley Additive exPlanations). The SHAP summary plot (Figure 4) highlights the importance and directionality of feature contributions. For example, the feature right_TemporalPole_original_glcm_Correlation_FA had the strongest and most consistent effect: low values increased the predicted probability of conversion, whereas high values decreased it. In contrast, right_FrontalOrbital Cortex_original_glszm_SmallAreaHighGrayLevelEmphasis_FA showed the opposite trend, where high values were associated with a higher probability of conversion.

**Figure 4.**
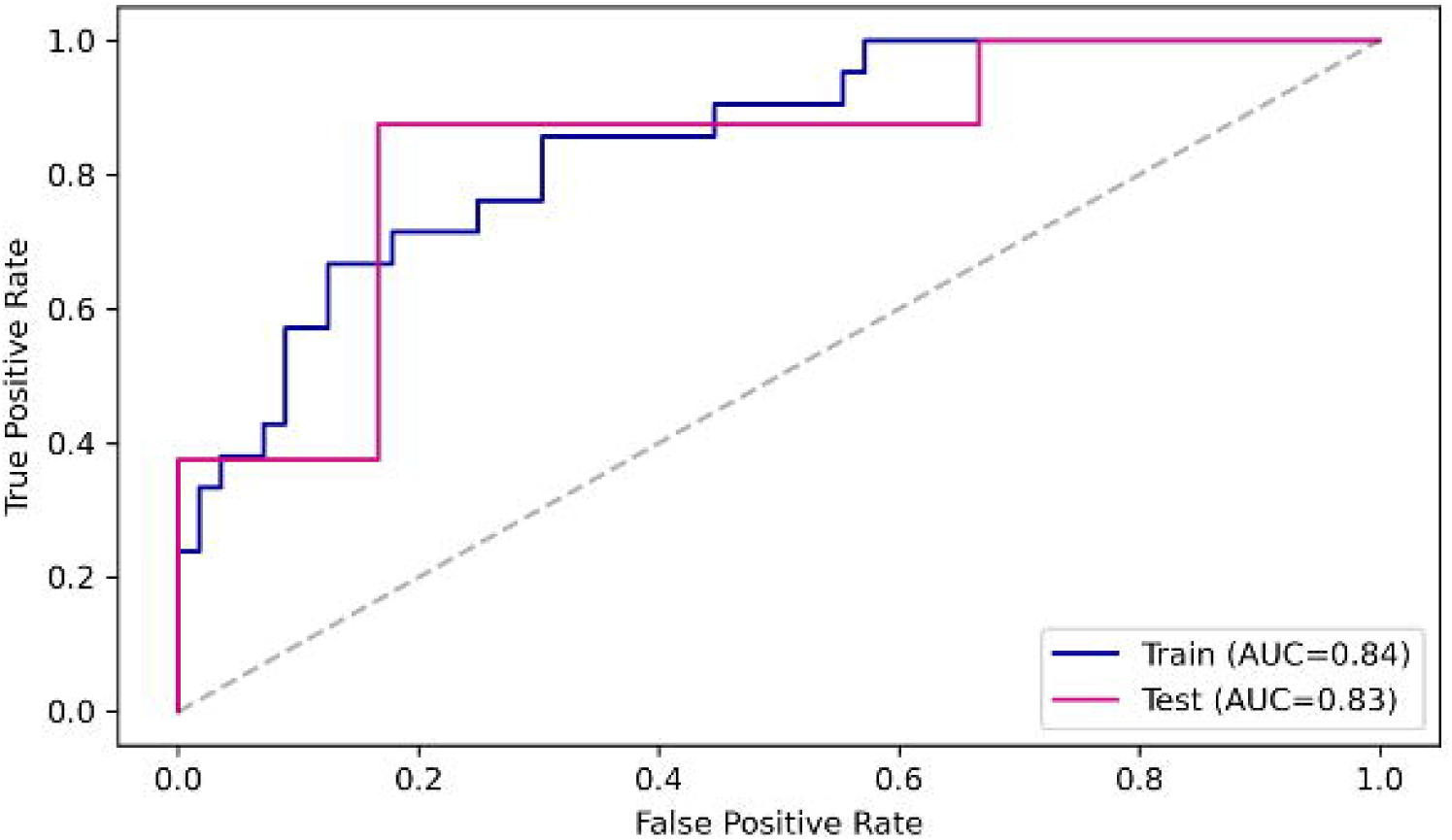
SHAP values illustrate each feature’s marginal contribution to the radiomics model output. · Topf-ranked features exhibit the most variability and strongest influence, consistent with model sparsity

Several features demonstrated non-linear effects or interactions, as indicated by mixed red/blue coloring on both sides of the SHAP axis. Features ranked lower had smaller SHAP value ranges, reflecting a less significant impact on the model’s output. Notably, selected features spanned both hemispheres and were derived from multiple texture matrices, including GLSZM, GLDM, GLCM, and first-order statistics, extracted from both FA and MD maps.

### Group Differences in Radiomic–Cognitive Correlations

To explore whether the relationships between selected radiomic features and cognitive scores differed between groups, we computed Pearson correlation coefficients separately for converter and non-converter groups. To assess whether the strength of association between radiomic features and cognitive scores differed between converters and non-converters, we conducted Fisher’s r-to-z transformation for each feature–score pair. Figure 5 visualizes the correlation values for each group. The analysis revealed three radiomic–cognition pairs with significantly different correlations between the two groups.

**Figure 5.**
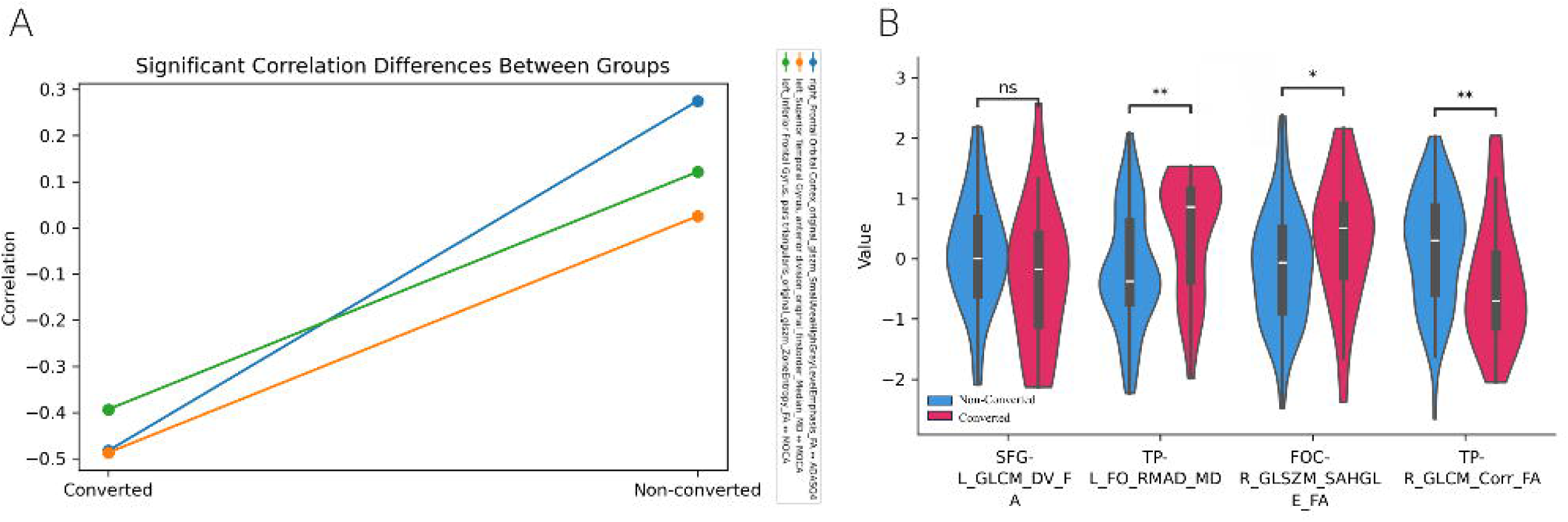
Differences in correlations and distributions of selected radiomic features between Converted and Non-Converted groups. · Line plot showing Pearson correlation coefficients between selected radiomic features and cognitive scores for each group. (B) Violin plots illustrating the distribution of the same features in Converted and Non-Converted individuals. Statistical differences between groups were assessed using independent two-sample t-tests or Mann–Whitney U tests, depending on normality. Significance levels are indicated above each feature (ns = not significant; * p < 0.05; ** p < 0.01). Abbreviations used in the figure: SFG-L_GLCM_DV_FA, left_Superior Frontal Gyrus_original_glcm_DifferenceVariance_FA; TP-L_FO_RMAD_MD, left_Temporal Pole_original_firstorder_RobustMeanAbsoluteDeviation_MD; FOC-R_GLSZM_SAHGLE_FA, right_Frontal Orbital Cortex_original_glszm_SmallAreaHighGrayLevelEmphasis_FA; TP-R_GLCM_Corr_FA, right_Temporal Pole_original_glcm_Correlation_FA

The feature right_Frontal Orbital Cortex_original_glszm_SmallAreaHighGrayLevelEmphasis_FA showed a positive correlation with ADASQ4 in the non-converted group (r = 0.274) but a strong negative correlation in the converted group (r = –0.483), with a statistically significant difference (p <0.0005). The feature left_Superior Temporal Gyrus, anterior division_original_firstorder_Median_MD had minimal correlation with MoCA in non-converters (r = 0.025) but a strong negative correlation in converters (r = –0.487), yielding a significant group difference (p = 0.016). Similarly, left_Inferior Frontal Gyrus, pars triangularis_original_glszm_ZoneEntropy_FA was weakly correlated with MoCA in non-converters (r = 0.121) but negatively correlated in converters (r = –0.39), also with a significant difference (p = 0.020). Interestingly, right_TemporalPole_original_glcm_Correlation_FA has a negative correlation with most cognitive tests (Table 3).

**Table 3.**
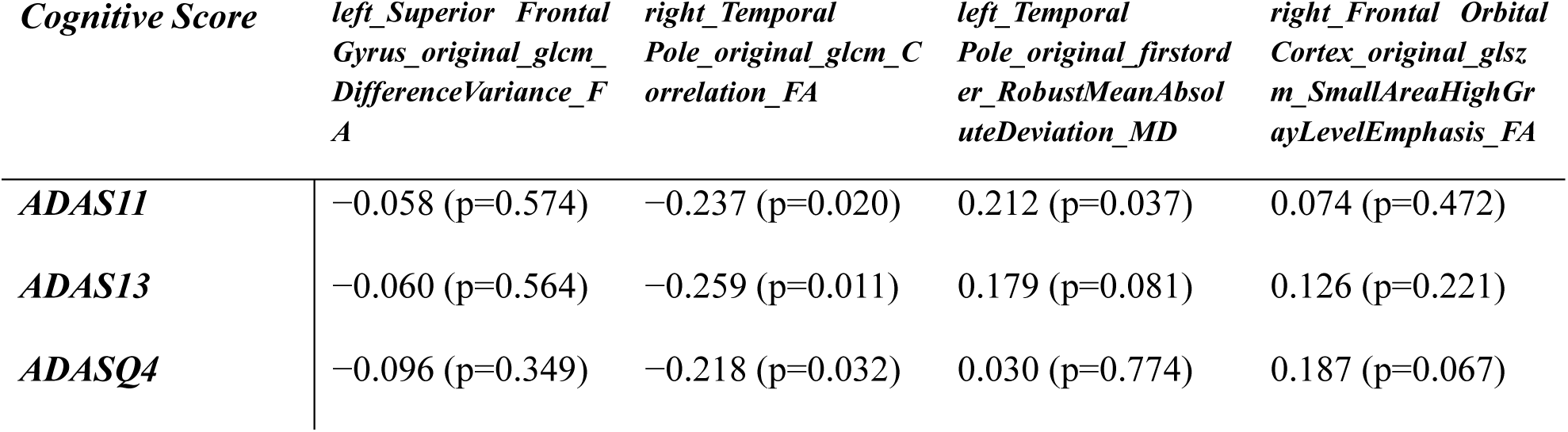
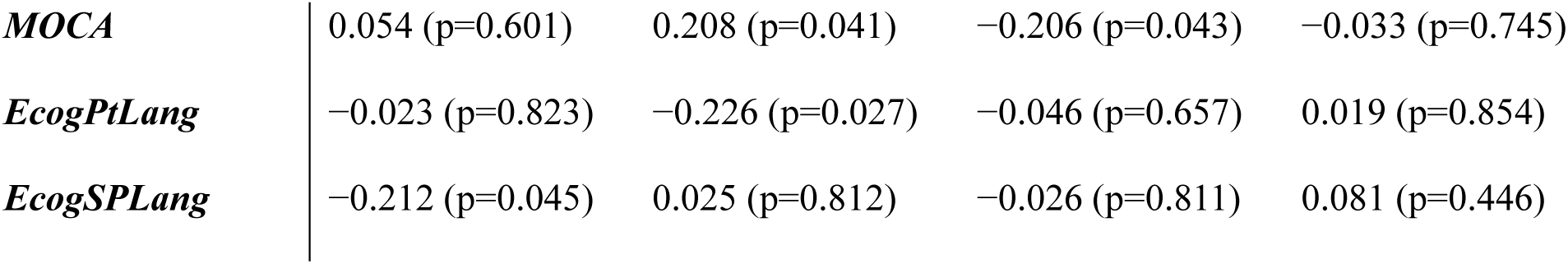
Pearson correlation coefficients (r) between selected diffusion-derived radiomic features (FA and MD) and cognitive scores, with corresponding p-values shown in parentheses.

These findings suggest that certain radiomic features are more closely associated with cognitive performance in individuals who convert to Alzheimer’s disease, indicating potential differences related to disease progression.

## Discussion

This study employed an interpretable radiomics framework to identify features distinguishing individuals with MCI who later converted to AD from those who did not. To our knowledge, this is the first investigation applying radiomics to DTI-derived FA and MD maps within gray matter regions of the language network to predict MCI to AD conversion. The resulting model demonstrated robust classification performance based on features extracted from fronto-temporal regions, underscoring the significance of the language network as a biomarker substrate for disease progression.

Using SHAP-based explainability, we found that the most influential discriminative features arose from frontal and temporal areas, particularly the temporal pole, superior temporal gyrus, inferior frontal gyrus, and orbitofrontal cortex. These findings align with extensive evidence linking fronto–temporal circuitry to neurodegeneration in AD and related disorders (Jin et al., 2024; Tang et al., 2024) Notably, the most predictive features were not limited to conventional diffusion metrics, FA and MD, but included higher-order texture descriptors derived from gray-level matrices. These describe spatial heterogeneity and organizational complexity within tissue, indicating that pathological changes in MCI converters manifest not only as global diffusivity alterations but also as disrupted microstructural patterning. This supports the notion that early AD pathology may involve cytoarchitectural disorganization in language hubs rather than uniform atrophy detectable by standard analyses.

Our results extend the DTI-radiomics literature, which has primarily focused on white matter tracts or non-DTI modalities (Cheng et al., 2023; Song et al., 2024; Zhao et al., 2020; Zhou et al., 2021). The present work expands upon prior research by applying radiomics analyses to cortical DTI-derived maps, providing interpretable biomarkers of preclinical alterations that are not detectable with conventional metrics. This approach aligns with prior evidence highlighting gray matter FA and MD abnormalities in disease progression (Marcos Dolado et al., 2019; Scola et al., 2010). Moreover, the focus on language-related areas is supported by documented early language impairments in individuals who later convert to AD (Liampas et al., 2023; Taler & Phillips, 2008) further, thereby addressing a critical gap in the existing literature (Ai et al., 2025). This approach offers an interpretable window into cortical microstructural vulnerability preceding cognitive symptoms.

Beyond classification, we examined how relationships between radiomic features and cognition diverged between converters and non-converters. Group-wise correlation comparisons revealed that microstructural–cognitive coupling was altered in converters, implicating network-specific dysregulation rather than uniform degradation. For instance, the SmallAreaHighGrayLevelEmphasis_FA feature in the right orbitofrontal cortex correlated positively with ADAS-Q4 performance in non-converters but negatively in converters. This suggests that microstructural complexity that supports function in healthy aging may signify pathology in those at risk for AD, reflecting a nonlinear or context-dependent shift in structure–function relationships. Similarly, increased MD in the left superior temporal gyrus was selectively associated with poorer MoCA performance in converters, while higher FA-entropy in the left inferior frontal gyrus corresponded to cognitive decline, both highlighting the vulnerability of language-related cortices to early neurodegenerative processes.

Collectively, these findings position language-network DTI radiomics as a promising tool for early risk stratification. By quantifying spatial heterogeneity beyond global FA or MD summaries, the framework enhances sensitivity to subtle microstructural pathology and facilitates biologically interpretable predictions. This approach may complement cognitive measures to improve personalized prediction models and enrich clinical trials targeting individuals at preclinical or prodromal stages of AD.

Nevertheless, certain limitations temper these conclusions. The modest sample size (n = 97; ADNI) restricts generalizability and raises potential overfitting concerns. The use of only FA and MD excludes advanced diffusion metrics (e.g., axial/radial diffusivity, NODDI), while the reliance on Harvard–Oxford parcellation limits subject-level anatomical precision. The absence of comprehensive language testing and the cross-sectional design constrain causal inference and longitudinal modeling. Future research should employ multi-site longitudinal datasets, incorporate multi-shell diffusion imaging, leverage individualized cortical parcellations, and integrate multimodal data, combining radiomics with clinical, neuropsychological, and functional measures, to refine predictive accuracy and interpretability for clinical translation.

## Conclusion

In summary, this study introduces a novel radiomics approach applying FA- and MD-based texture analysis within gray matter language networks to predict conversion from MCI to AD. The findings highlight the diagnostic relevance of fronto-temporal microstructural heterogeneity and provide interpretable, biologically grounded markers of early neurodegenerative change. These results pave the way for integrative, multimodal predictive frameworks combining diffusion-based radiomics with cognitive and functional data to advance individualized risk profiling and early therapeutic targeting in Alzheimer’s disease.

## Data Availability

All data produced in the present study are available upon reasonable request to the authors

## Declarations

### Ethics approval and consent to participate

The ADNI study was conducted according to the Good Clinical Practice guidelines, the Declaration of Helsinki, and U.S. 21 CFR Part 50 (Protection of Human Subjects), and approved by the Institutional Review Boards of all participating institutions. Written informed consent was obtained from all participants or authorized representatives before their inclusion in ADNI.

### Consent for publication

Not applicable

### Availability of data and materials

The datasets used and/or analysed during the current study are available from the corresponding author on reasonable request.

### Authors’ contributions

SR proposed the initial research idea and supervised the study. FJ contributed to study design, performed radiomics feature extraction, machine learning modeling, and statistical analyses, and coordinated the research workflow. MH, MK, FZ, and MS contributed to data organization, preprocessing, and neuroimaging data processing, and participated in writing different sections of the manuscript. FJ drafted the main version of the manuscript. All authors contributed to manuscript revision and approved the final version.

## Acknowledgements

The authors would like to thank the Interdisciplinary Schools and Moslem Solhirad for their support. We acknowledge ADNI investigators and participants for providing the data used in this study. We are also grateful to Dr. Arash Zare Sadeghi for his valuable assistance.

## References

Ai, M., Liu, Y., Liu, D., Yan, C., Wang, X., & Chen, X. (2025). Research progress in predicting the conversion from mild cognitive impairment to Alzheimer’s disease via multimodal MRI and artificial intelligence [Mini Review]. Frontiers in Neurology, Volume 16- 2025. 10.3389/fneur.2025.1596632

Bruggen, T. v., Stieltjes, B., Thomann, P. A., Parzer, P., Meinzer, H.-P., & Fritzsche, K. H. (2012). Do Alzheimer-specific microstructural changes in mild cognitive impairment predict conversion? Psychiatry Research: Neuroimaging, 203, 184–193.

Chandra, A., Dervenoulas, G., Politis, M., & Initiative, A. s. D. N. (2019). Magnetic resonance imaging in Alzheimer’s disease and mild cognitive impairment. Journal of neurology, 266, 1293–1302.

Cheng, X., Li, D., Peng, J., Shu, Z., & Xing, X. (2023). Prediction of Mild Cognitive Impairment Progression to Alzheimer’s Disease Based on Diffusion Tensor Imaging-Derived Diffusion Parameters: Construction and Validation of a Nomogram. Eur Neurol, 86(6), 408–417. 10.1159/000534767

Desikan, R. S., Ségonne, F., Fischl, B., Quinn, B. T., Dickerson, B. C., Blacker, D., Buckner, R. L., Dale, A. M., Maguire, R. P., Hyman, B. T., Albert, M. S., & Killiany, R. J. (2006). An automated labeling system for subdividing the human cerebral cortex on MRI scans into gyral based regions of interest. Neuroimage, 31(3), 968–980. 10.1016/j.neuroimage.2006.01.021

Jin, S., Wang, J., & He, Y. (2024). The brain network hub degeneration in Alzheimer’s disease. Biophys Rep, 10(4), 213–229. 10.52601/bpr.2024.230025

Kumar, V., Gu, Y., Basu, S., Berglund, A., Eschrich, S. A., Schabath, M. B., Forster, K., Aerts, H. J., Dekker, A., & Fenstermacher, D. (2012). Radiomics: the process and the challenges. Magnetic resonance imaging, 30(9), 1234–1248.

Liampas, I., Folia, V., Morfakidou, R., Siokas, V., Yannakoulia, M., Sakka, P., Scarmeas, N., Hadjigeorgiou, G., Dardiotis, E., & Kosmidis, M. H. (2023). Language Differences Among Individuals with Normal Cognition, Amnestic and Non-Amnestic MCI, and Alzheimer’s Disease. Arch Clin Neuropsychol, 38(4), 525–536. 10.1093/arclin/acac080

Marcos Dolado, A., Gomez-Fernandez, C., Yus Fuertes, M., Barabash Bustelo, A., Marcos-Arribas, L., Lopez-Mico, C., Jorquera Moya, M., Fernandez-Perez, C., Montejo Carrasco, P., Cabranes Diaz, J. A., Arrazola Garcia, J., & Maestu Unturbe, F. (2019). Diffusion Tensor Imaging Measures of Brain Connectivity for the Early Diagnosis of Alzheimer’s Disease. Brain Connect, 9(8), 594–603. 10.1089/brain.2018.0635

Scola, E., Bozzali, M., Agosta, F., Magnani, G., Franceschi, M., Sormani, M. P., Cercignani, M., Pagani, E., Falautano, M., Filippi, M., & Falini, A. (2010). A diffusion tensor MRI study of patients with MCI and AD with a 2-year clinical follow-up. J Neurol Neurosurg Psychiatry, 81(7), 798–805. 10.1136/jnnp.2009.189639

Skolariki, K., Terrera, G. M., & Danso, S. O. (2021). Predictive models for mild cognitive impairment to Alzheimer’s disease conversion. Neural Regeneration Research, 16(9), 1766–1767. 10.4103/1673-5374.306071

Song, Q., Peng, J., Shu, Z., Xu, Y., Shao, Y., Yu, W., & Yu, L. (2024). Predicting Alzheimer’s progression in MCI: a DTI-based white matter network model. BMC Medical Imaging, 24(1), 103. 10.1186/s12880-024-01284-7

Taler, V., & Phillips, N. A. (2008). Language performance in Alzheimer’s disease and mild cognitive impairment: a comparative review. J Clin Exp Neuropsychol, 30(5), 501–556. 10.1080/13803390701550128

Tang, X., Guo, Z., Chen, G., Sun, S., Xiao, S., Chen, P., Tang, G., Huang, L., & Wang, Y. (2024). A Multimodal Meta-Analytical Evidence of Functional and Structural Brain Abnormalities Across Alzheimer’s Disease Spectrum. Ageing Res Rev, 95, 102240. 10.1016/j.arr.2024.102240

van Griethuysen, J. J. M., Fedorov, A., Parmar, C., Hosny, A., Aucoin, N., Narayan, V., Beets-Tan, R. G. H., Fillion-Robin, J.-C., Pieper, S., & Aerts, H. J. W. L. (2017). Computational Radiomics System to Decode the Radiographic Phenotype. Cancer Research, 77(21), e104–e107. 10.1158/0008-5472.Can-17-0339

Wang, Y., Li, Q., Yao, L., He, N., Tang, Y., Chen, L., Long, F., Chen, Y., Kemp, G. J., Lui, S., & Li, F. (2024). Shared and differing functional connectivity abnormalities of the default mode network in mild cognitive impairment and Alzheimer’s disease. Cerebral Cortex, 34(3). 10.1093/cercor/bhae094

Zhao, K., Ding, Y., Han, Y., Fan, Y., Alexander-Bloch, A. F., Han, T., Jin, D., Liu, B., Lu, J., & Song, C. (2020). Independent and reproducible hippocampal radiomic biomarkers for multisite Alzheimer’s disease: diagnosis, longitudinal progress and biological basis. Science Bulletin, 65(13), 1103–1113.

Zhou, H., Jiang, J., Lu, J., Wang, M., Zhang, H., Zuo, C., & , A. s. D. N. I. (2019). Dual-Model Radiomic Biomarkers Predict Development of Mild Cognitive Impairment Progression to Alzheimer’s Disease [Original Research]. Frontiers in Neuroscience, Volume 12 - 2018. 10.3389/fnins.2018.01045

Zhou, P., Zeng, R., Yu, L., Feng, Y., Chen, C., Li, F., Liu, Y., Huang, Y., & Huang, Z. (2021). Deep-Learning Radiomics for Discrimination Conversion of Alzheimer’s Disease in Patients With Mild Cognitive Impairment: A Study Based on (18)F-FDG PET Imaging. Front Aging Neurosci, 13, 764872. 10.3389/fnagi.2021.764872

